# Identification and validation of biomarkers *FAM107A*, *RHOBTB1* and *ZBTB16* associated with dietary restriction in osteoarthritis

**DOI:** 10.1101/2025.11.20.25340682

**Authors:** Siyuan Wen, Xuan Li, Zhongmao Xu, Junfei Wang

## Abstract

**Objective:** Osteoarthritis (OA), a chronic ailment that leads to joint degeneration, could be prevented by dietary restriction (DR). This study investigated the molecular mechanisms related to DR-related genes (DRRGs) as potential biomarkers in OA.

**Methods:** Transcriptome data of OA (GSE55235 and GSE89408) were retrieved from public databases. Differentially expressed genes (DEGs) from GSE55235 were cross-referenced with DRRGs to identify candidate genes. Machine learning techniques, expression validation, and receiver operating characteristic (ROC) curve analysis were utilized to pick out biomarkers. Functional enrichment analysis explored associated pathways, and the CIBERSORT algorithm assessed immune cell infiltration. Subsequently, an exploration was made into the associations between biomarkers and immune cells. Additionally, small-molecule compounds for treating OA were predicted and validated based on the biomarkers, and the magnitudes of biomarker expression were verified.

**Results:** A sum of 43 DR-related candidate genes in OA was determined. Among them, *FAM107A*, *RHOBTB1*, and *ZBTB16* were identified as biomarkers with strong predictive value for OA (AUC ≥ 0.7). Pathway enrichment analysis indicated that 3 biomarkers were significantly connected with the lysosome pathway. In terms of immune infiltration analysis, it revealed that the highest degree of positive correlation (r =0.72, *P* < 0.001) existed between *FAM107A* and CD8 T cells. Meanwhile, the most pronounced negative correlation (r=-0.74, *P* < 0.001) was found between *RHOBTB1* and activated mast cells. Moreover, molecular docking analysis demonstrated that 4-(5-benzo(1,3) dioxol-5-yl-4-pyridin-2-yl-1H-imidazol-2-yl) benzamide, benzo(a)pyrene, and tetrachlorodibenzodioxin had good binding affinities with the biomarkers. In addition, Reverse transcription quantitative polymerase chain reaction (RT-qPCR) confirmed that, in the OA group, the expression levels of *RHOBTB1* and *ZBTB16* increased, whereas the level of *FAM107A* was decreased (*P* < 0.05).

**Conclusion:** This research identified 3 DR-related biomarkers in OA, emphasizing that they were involved in the pathogenesis of OA and had the potential to serve as therapeutic targets.

## 1 Introduction

Osteoarthritis (OA) is the most prevalent degenerative joint disease and a major cause of disability worldwide. It is characterized by articular cartilage degeneration, synovial inflammation, osteophyte formation, and subchondral bone remodeling, leading to pain, stiffness, and impaired joint function, particularly in the elderly population[1–3]. The increasing prevalence of OA, driven by aging, obesity, and mechanical stress, has imposed heavy socioeconomic and health burdens[4, 5]. OA is a highly prevalent joint disorder and a major contributor to disability worldwide. Despite its burden, the diagnosis of OA in clinical practice still predominantly depends on symptom assessment and conventional imaging modalities such as radiography and magnetic resonance imaging (MRI). These methods, however, demonstrate important limitations. Radiographic joint space narrowing, although widely adopted as the standard endpoint for disease-modifying osteoarthritis drug (DMOAD) trials, is only an indirect marker of cartilage health and lacks sufficient sensitivity and specificity to detect early degenerative changes[6]. As a result, subtle structural and biochemical alterations in articular cartilage often remain undetected, leading to diagnostic delays and inconsistent clinical outcomes.

In recent years, increasing attention has been directed toward the concept of early- or pre-osteoarthritis, which highlights the importance of timely detection before irreversible joint damage occurs. Advances in imaging, particularly quantitative and compositional MRI techniques, have enabled visualization of early pathological changes across cartilage, bone marrow, menisci, and synovium[7]. These developments underscore a paradigm shift from palliative care of late-stage OA toward early diagnosis and intervention, with the potential to improve patient outcomes and enhance the success of disease-modifying strategies.

Although various therapeutic approaches for symptom management are currently available, including pharmacological interventions, physical therapy, and surgical procedures, existing treatment strategies are primarily limited to symptom relief and cannot fundamentally halt or reverse the pathological progression of osteoarthritis. This current situation highlights the inadequacy of present treatment paradigms and underscores the urgent need for in-depth exploration of the molecular pathogenesis of osteoarthritis and the development of disease-modifying therapeutic strategies[8].

Recent research has revealed that OA is not merely a mechanical “wear and tear” disorder but also involves complex molecular and immunometabolic pathways. Aberrant activation of signaling cascades such as Indian hedgehog (Ihh), immune dysregulation including indoleamine 2,3-dioxygenase 1 (IDO1) overexpression, ferroptosis-related gene expression, and immune cell infiltration all contribute to chondrocyte hypertrophy, extracellular matrix breakdown, and disease progression[1, 5, 9]. Bioinformatics-driven approaches have identified OA-associated biomarkers and inflammatory infiltrates, providing valuable insights into disease pathogenesis and potential diagnostic targets[4, 9].

Meanwhile, dietary restriction (DR), defined as reduced caloric intake without malnutrition, has emerged as the most effective non-genetic intervention to extend healthspan and lifespan across multiple species[10]. DR influences nutrient-sensing pathways such as insulin/IGF-1, mTOR, AMPK, and sirtuins, thereby reducing oxidative stress, improving mitochondrial function, and enhancing autophagy[5, 10]. In addition, DR modulates systemic and local immune responses, reshapes gut microbiota, and exerts protective effects against age-related diseases including metabolic disorders, neurodegeneration, and cancer[5, 8]. Although DR has been widely studied in other contexts, its specific molecular roles in OA pathogenesis remain unclear.Dietary restriction has recently gained increasing attention in OA research, with two main approaches being caloric restriction and intermittent fasting. Animal studies have shown that caloric restriction using standard chow can delay OA onset, whereas this protective effect is attenuated under a high-fat diet, likely due to elevated systemic inflammation[11]. Clinical evidence from a systematic review and meta-analysis of randomized trials demonstrated that dietary interventions significantly improve pain, physical function, and body weight in OA patients, with reduced energy diets showing the greatest effectiveness[12]. Intermittent fasting, characterized by cyclic caloric restriction and metabolic switching, exerts multi-target protective effects by improving metabolic markers, reducing inflammation, activating autophagy, and modulating gut microbiota[13]. Despite these promising findings, the precise molecular mechanisms, particularly the role of dietary restriction–related genes in OA pathogenesis, remain unclear. Elucidating these mechanisms is crucial for developing precise non-pharmacological intervention strategies. Integrating evidence from molecular biology, immunology, and metabolism, exploring the potential links between DR-related molecular activities and OA development may provide new perspectives for identifying biomarkers and designing non-pharmacological interventions. This study therefore aims to investigate the interaction between dietary restriction and osteoarthritis, with the goal of uncovering novel molecular targets that could guide future diagnostic and therapeutic strategies.

## 2 Materials and methods

### 2.1 Retrieval and collection of gene expression data

Transcriptome datasets linked to osteoarthritis (OA) were retrieved from the Gene Expression Omnibus database (GEO). Specifically, the GSE55235 was established on the GPL96 platform. It consisted of 10 synovial tissue specimens from OA patients (OA group) and 10 synovial tissue specimens from healthy individuals (control group), and this dataset was used as the training set[14]. The GSE89408 dataset, on the other hand, was high-throughput transcriptome sequencing data. Based on the GPL11154 platform, it included 22 synovial tissue samples from OA patients (OA group) and 28 synovial tissue samples from normal controls (control group), which served as the validation set[14]. Additionally, a collection of 276 dietary restriction-related genes (DRRGs) was retrieved from the Dietary Restriction Gene database (GenDR)[9] **(S1 Table)**.

### 2.2 Discovery of differentially expressed genes (DEGs)

To uncover the genes with DEGs between the OA and control individuals within the training set, the R package "limma" (v 3.56.4)[15] was utilized. The selection criteria for these DEGs were |log_2_ Fold Change (FC)| > 0.5 and a *P*-value (*P*) < 0.05. Moreover, the "ggplot2" (v 3.5.1) R package[16] was utilized to construct volcano plots of the top 10 up-regulated/down-regulated DEGs, and the "ComplexHeatmap" R package (v 2.18.0)[17] was used to generate heatmaps of the top 50 up-regulated/down-regulated DEGs.

### 2.3 Acquisition and analysis of candidate genes

The "VennDiagram" R package (v 1.7.3)[18] was utilized to obtain candidate genes by intersecting DEGs with DRRGs. The "clusterProfiler" R package (v 4.15.0.003)[19] was employed to conduct GO and KEGG enrichment analyses on the obtained candidate genes (adj.*P* < 0.05). For investigating the protein-level interactions of the candidate genes, the STRING database was employed.

Specifically, the candidate genes were fed into this database, and genes having an interaction score > 0.4 were picked out. Subsequently, Cytoscape (v 3.10.3)[20] was applied for the construction of a PPI network.

### 2.4 Characterized gene selection via machine learning

The "randomForest" R package (v 4.7-1.2)[21] was applied to construct a random forest classification model relying on candidate genes. By evaluating the importance scores of each candidate gene, RF genes were obtained. Subsequently, the "glmnet" R package (v4.1-8) (https://glmnet.stanford.edu) was used to conduct LASSO analysis on the RF genes. Key parameters were set as standardize=True, alpha=1, and family=family. Through the implementation of 10-fold cross-validation, the optimal lambda value was established to define the characterized genes.

### 2.5 Determining Biomarkers for OA

The Wilcoxon test was utilized to assess disparities in the expression of characterized genes between the OA and control individuals within both validation and training sets (*P* < 0.05). Characterized genes that exhibited remarkable expression differences and consistent patterns of change across 2 datasets were defined as candidate biomarkers (*P* < 0.05). Subsequently, in all samples from 2 datasets, the R package "pROC" (v 1.18.5)[22] was employed to evaluate the candidate biomarkers. The ROC curve was plotted to analyze their ability to distinguish between the OA and control individuals. The AUC value was calculated, and candidate biomarkers with an AUC ≥ 0.7 in both datasets were determined as biomarkers.

### 2.6 Gene Interaction and localization analysis of biomarkers

The GeneMANIA database was employed to predict genes associated with the functions of biomarkers and construct a gene-gene interaction (GGI) network. The "Rcircos" R package (v 1.2.2)[23] was utilized to analyze the distribution of biomarkers on human chromosomes. Additionally, the target protein sequence files corresponding to the biomarkers were obtained from the NCBI official website. Subsequently, leveraging the target protein sequences, the DeepLoc 2.0 was utilized to forecast the subcellular distribution details of the biomarkers.

### 2.7 GSEA

The R package "clusterProfiler" (v 4.15.0.003) was applied to execute GSEA to identify the signaling pathways associated with the biomarkers. Additionally, for the training set, the "psych" package (v 2.4.6.26)[24] was utilized to carry out a Spearman correlation analysis. The relationship between biomarkers and all other genes in both the OA and control individuals. Genes were ranked in descending order based on their Spearman correlation coefficients, producing a list of related genes for each biomarker. For the GSEA analysis, the "c2.cp.kegg.v7.5.1.symbols.gmt" was obtained from the MSigDB. The screening criteria included an |NES| > 1, a *P* < 0.05, and an FDR < 0.25. These outcomes were graphically shown by making use of the "enrichplot" package (v 1.18.3)[24]. Specifically, the top 5 signaling pathways were presented, ranked according to their *P*-values.

### 2.8 Immune infiltration analysis

Within the training set, the CIBERSORT algorithm was utilized to calculate the relative abundances and proportions of immune infiltration for 22 types of immune cells in each sample of the OA and control group[25]. The Wilcoxon test was utilized to assess the disparities in the proportions of 22 immune cell types across 2 groups (*P* < 0.05). For the visual representation of the aforesaid results, the R package "ggplot2" (v 3.5.1) was employed. Subsequently, the "psych" R package (v 2.4.6.26) was applied to conduct Spearman correlation analyses. These analyses were performed to examine the relationships: one was between immune cells with differential abundances, and the other was between these differentially abundant immune cells and biomarkers (*P* < 0.05, |correlation (r)| > 0.3). The "corrplot" R package (v 0.95)[25] was used to generate correlation heatmaps.

### 2.9 Molecular regulatory network

To delve into the regulatory mechanisms involved with biomarkers. The miRDB database was utilized to predict the interaction pairs between miRNAs and messenger RNAs, including the gene transcripts associated with biomarkers. Subsequently, miRNAs corresponding to each biomarker were retrieved, and a regulatory network about miRNA-mRNA was built. The ChEA3 database was employed to predict TFs associated with biomarkers. The top 30 TFs were selected based on their score and used to construct a TF-mRNA regulatory network. Both regulatory networks were constructed using Cytoscape (v 3.10.3).

### 2.10 Small molecule compound prediction and molecular docking for exploring potential therapeutic agents in OA

To identify potential therapeutic drugs or small-molecule compounds associated with biomarkers in OA, the CTD was searched, and relevant small-molecule compound information was extracted. The top 20 potential small-molecule compounds for each biomarker were selected based on the Interaction count. The interaction network between these small-molecule compounds and biomarkers was visualized using Cytoscape (v 3.10.3) software. To further investigate the binding ability between biomarkers and the top 3 targeted small molecule compounds ranked by interaction count, molecular docking analysis was performed. The protein structures of biomarkers were obtained from the AlphaFold DB Database, and the 3-dimensional structures of targeted small molecule compounds were downloaded from the PubChem database. Subsequently, Autodock software (v 4.0)[26] was employed to perform a molecular docking analysis between the biomarkers and the key targeted small-molecule compounds. Additionally, the binding energy of the molecular docking was computed. A binding energy < -5.0 kcal/mol was regarded as suggesting satisfactory binding activity. PyMOL software (v 3.0.3)[27] was applied to assess the binding activity between the small molecule compounds and the targets according to the docking energy values.

### 2.11 RT-qPCR

During the course of this research, 5 synovial tissue specimens were procured from patients with OA. Simultaneously, an additional 5 were obtained from healthy control individuals at Affiliated Drum Tower Hospital, Medical School of Nanjing University. Subsequently, these samples were applied to perform RT-qPCR analysis. Ethical approval for this research was obtained from 2025-0615-01. The TRIzol reagent (Ambion, USA) was used to extract total RNA from the 10 tissue samples. The concentration of the isolated RNA was measured using a NanoPhotometer N50. Subsequently, reverse transcription was performed to generate cDNA. This was achieved by using the SureScript First-Strand cDNA Synthesis Kit in combination with an S1000TM Thermal Cycler (Bio-Rad, USA). The primer sequences applied for qPCR were presented in **S2 Table**. The quantitative PCR was conducted on a CFX Connect quantitative instrument (Bio-Rad, USA). The 2^-ΔΔCT^ method was adopted to determine the relative quantification of prognostic genes. The RT-qPCR data were analyzed and graphically displayed by GraphPad Prism 10 software (*P* < 0.05).

### 2.12 Statistical analysis

R software (v 4.2.2) was utilized to accomplish all bioinformatics analyses. For the comparison among different groups, the Wilcoxon test was utilized. Unless otherwise indicated, statistical significance was established when *P* < 0.05. In the context of RT-qPCR analysis, inter-group comparisons were carried out through the application of the t-test (*P* < 0.05).

## 3 Results

### 3.1 Discovery, GO/KEGG analysis, and PPI network construction of candidate genes

In the training set of this research, a compilation of 1,451 DEGs was successfully identified between the OA and control groups. Particularly, in contrast to the control group, 754 genes were found to be up-regulated and 697 genes were noted as down-regulated in the OA group. The distribution differences of these DEGs were visually presented through volcano plots and heatmaps (**Fig 1A-B**). By taking the intersection of 276 DRRGs and 1,451 DEGs, 43 candidate genes were successfully obtained **(S3 Table, Fig 1C**). The GO analysis of these 43 candidate genes clearly revealed their enriched biological processes. Specifically, it covered 10 cellular components, 233 biological processes, and 24 molecular functions (adj.*P* < 0.05) **(S4 Table, Fig 1D**). In terms of biological processes, the candidate genes were significantly enriched in functions such as glucose homeostasis, carbohydrate homeostasis, response to insulin, rhythmic process, and response to peptide hormone. From the perspective of cellular components, these genes were significantly associated with the membrane raft, membrane microdomain, protein kinase complex, leading edge membrane, and caveola. At the molecular functions level, the functions linked to candidate genes predominantly encompassed phosphatidylinositol 3-kinase binding, insulin receptor binding, dipeptidyl-peptidase activity, the alcohol dehydrogenase [NAD(P)+] activity, and insulin-like growth factor receptor I binding. Furthermore, the KEGG pathway analysis results showed that these 43 candidate genes were highly concentrated in 20 pathways, including the FoxO signaling pathway, the longevity regulating pathway, and longevity regulating pathway-multiple species (adj. *P* < 0.05) **(S5 Table, Fig 1E**). A PPI network was constructed, which consisted of 29 proteins and encompassed 48 pairs of interaction relationships (**Fig 1F**).

**Fig 1.**
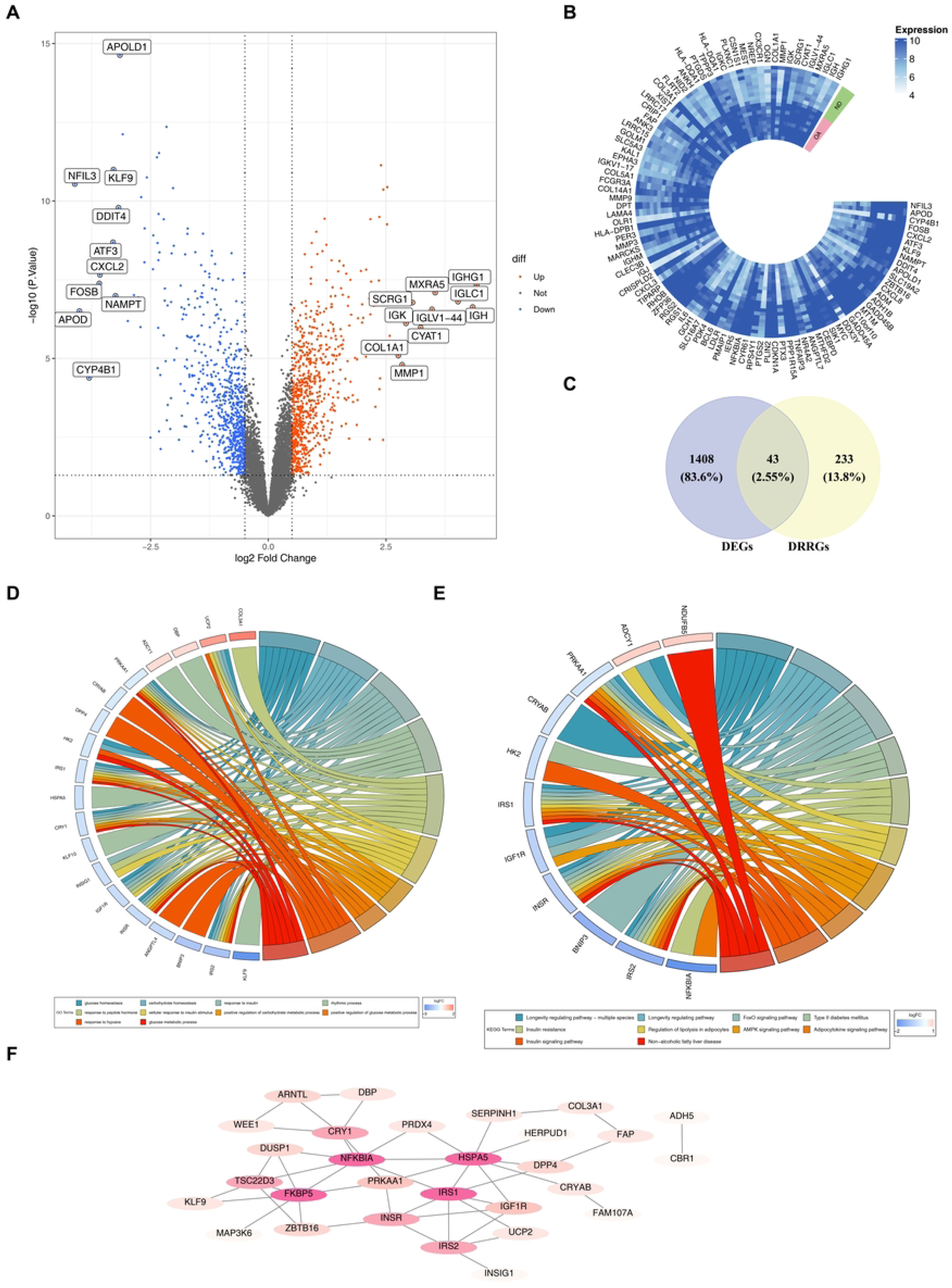
Identification and analyses of differentially expressed genes (DEGs) and candidate genes. (A) Volcano plot illustrating the distribution of DEGs between osteoarthritis (OA) and control groups. Red dots represent up-regulated genes, blue dots represent down-regulated genes, and labeled genes are of particular interest. (B) Circular heatmap showing the expression patterns of DEGs between OA and control groups. The color scale indicates the expression levels, with different clusters of genes visualized in a circular layout. (C) Venn diagram depicting the intersection of DEGs and diet-restriction related genes (DRRGs). A total of 43 candidate genes were identified at the intersection, with 1408 DEGs unique to DEGs and 233 DRRGs unique to DRRGs. (D) Circular chord diagram illustrating the enrichment of 43 candidate genes in KEGG pathways. The connections represent the associations between genes and pathways, with different colors distinguishing various pathways and genes. (E) Circular chord diagram highlighting the interactions involved in key signaling pathways related to the 43 candidate genes. The chords depict the relationships between genes and pathway components. (F) Protein-protein interaction (PPI) network of the 43 candidate genes. Nodes represent proteins, and edges represent interaction relationships, showing a network consisting of 29 proteins and 48 interaction pairs.

### 3.2 A sum of 6 characterized genes was obtained

RF was employed to screen the top 10 candidate genes based on their importance, namely *DUSP1*, *RHOBTB1*, *FAM107A*, *ZBTB16*, *WEE1*, *BNIP*3, *DBP*, *NFKBIA*, *UCP2*, and *IRS2* (**Fig 2A-B**). Subsequently, LASSO regression was utilized to further simplify the RF genes. When the optimal lambda values were reached, with log(lambda. min)=-7.56 and log(lambda 1se)=-6.91, a sum of 6 characteristic genes was recognized (**Fig 2C-D**), including *FAM107A*, *ZBTB16*, *IRS2*, *NFKBIA*, *RHOBTB1*, and *UCP2*.

**Fig 2.**
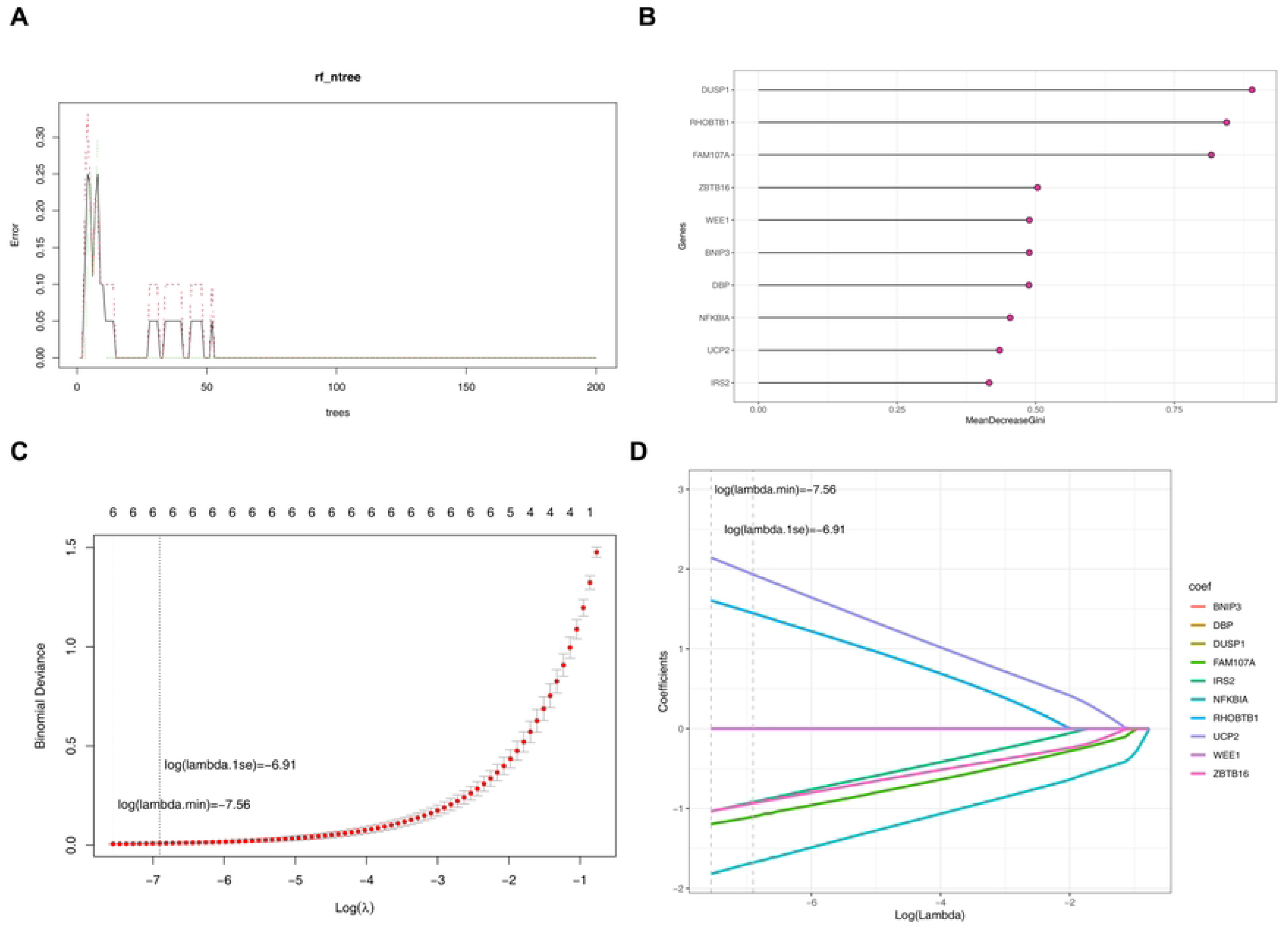
Identification of characterized genes. (A) Rf_ntree plot illustrating the error variation with the number of trees in the Random Forest model during the screening of candidate genes. (B) Mean Decrease Gini plot showing the top 10 candidate genes ranked by their importance in the Random Forest analysis: *DUSP1*, *RHOBTB1*, *FAM107A*, *ZBTB16*, *WEE1*, *BNIP3*, *DBP*, *NFKBIA*, *UCP2*, and *IRS2*. (C) LASSO regression plot of binomial deviance against log(λ), indicating the optimal λ values (log(lambda.min) = -7.56) for feature selection. (D) LASSO coefficient profile plot demonstrating the shrinkage of coefficients for the selected genes at the optimal λ values.

### 3.3 Identification of *FAM107A*, *RHOBTB1*, and *ZBTB16* as biomarkers

Within the training and validation datasets, significant and consistent disparities in the expression levels of the identified genes *FAM107A*, *RHOBTB1*, and *ZBTB16* were detected between the OA and control individuals (*P* < 0.05) (**Fig 3A**). Subsequently, additional ROC analysis, when performed across all samples in the 2 datasets, revealed that the AUC ≥ 0.7 for these 3 genes (**Fig 3B-C**). These results indicated that *FAM107A*, *RHOBTB1*, and *ZBTB16* exhibit a satisfactory ability to discriminate between OA and control samples, suggesting their potential diagnostic value. Consequently, *FAM107A*, *RHOBTB1*, and *ZBTB16* were identified as biomarkers for osteoarthritis.

**Fig 3.**
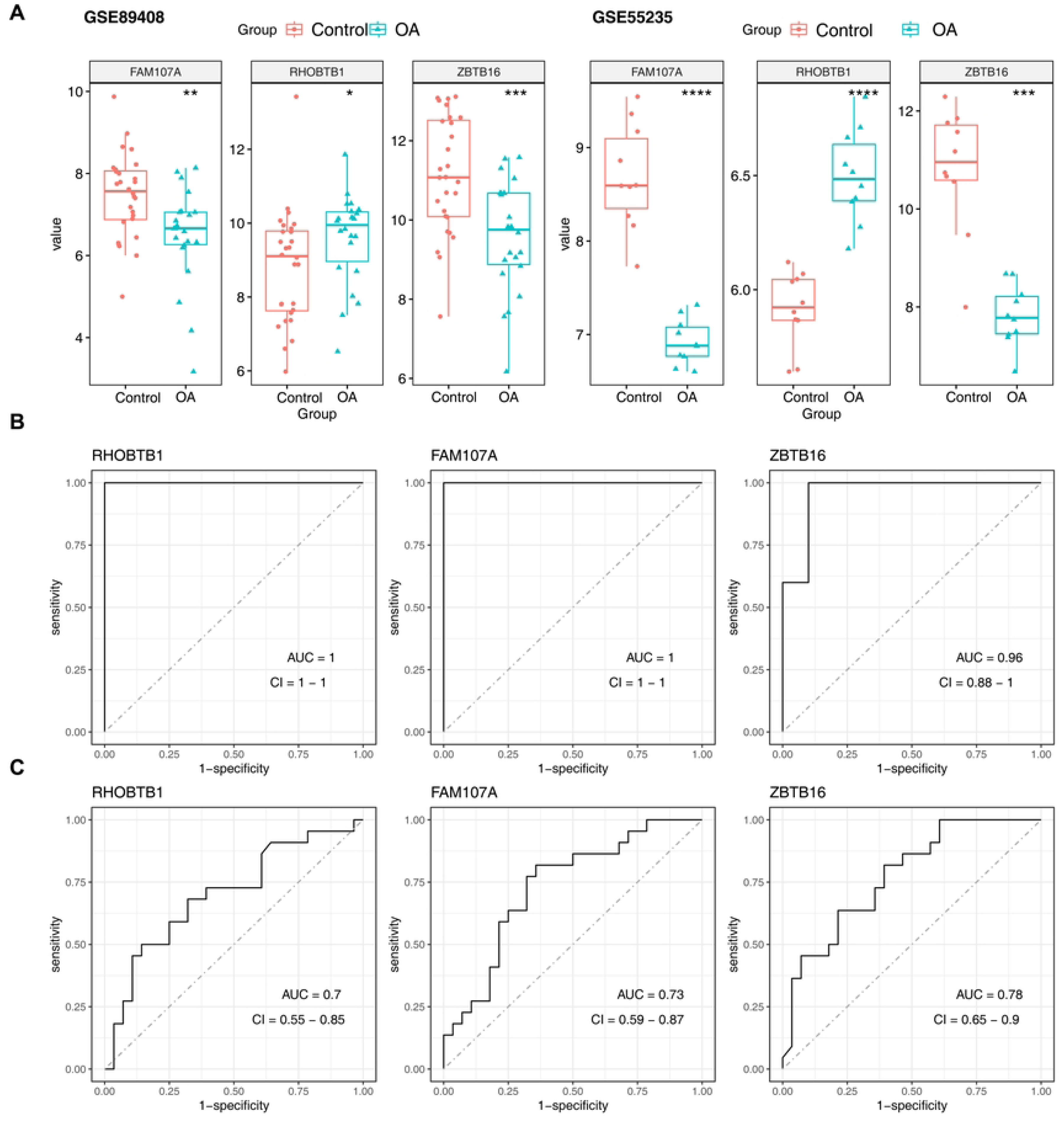
Identification of biomarkers. (A) Validation of biomarker expression levels. (B) Receiver operating characteristic analysis (ROC) analysis of biomarkers in the training set. (C) ROC analysis of biomarkers in the validation set.

### 3.4 Analysis of genes related to 3 biomarkers

A total of 20 functionally-related genes were predicted by the 3 biomarkers, including *FAM107B*, *CUL3*, *RUNX1T1*, etc. These genes were associated with functions such as transcription regulator complex and response to steroid hormone (**Fig 4A**). Chromosomal localization analysis revealed that *FAM107A*, *RHOBTB1*, and *ZBTB16* were located on chromosomes 3, 10, and 11, respectively (**Fig 4B**). Subcellular distribution analysis indicated that both *FAM107A* and *RHOBTB1* were located in the cytoplasm, whereas *ZBTB16* was situated in the nucleus (**Fig 4C**).

**Fig 4.**
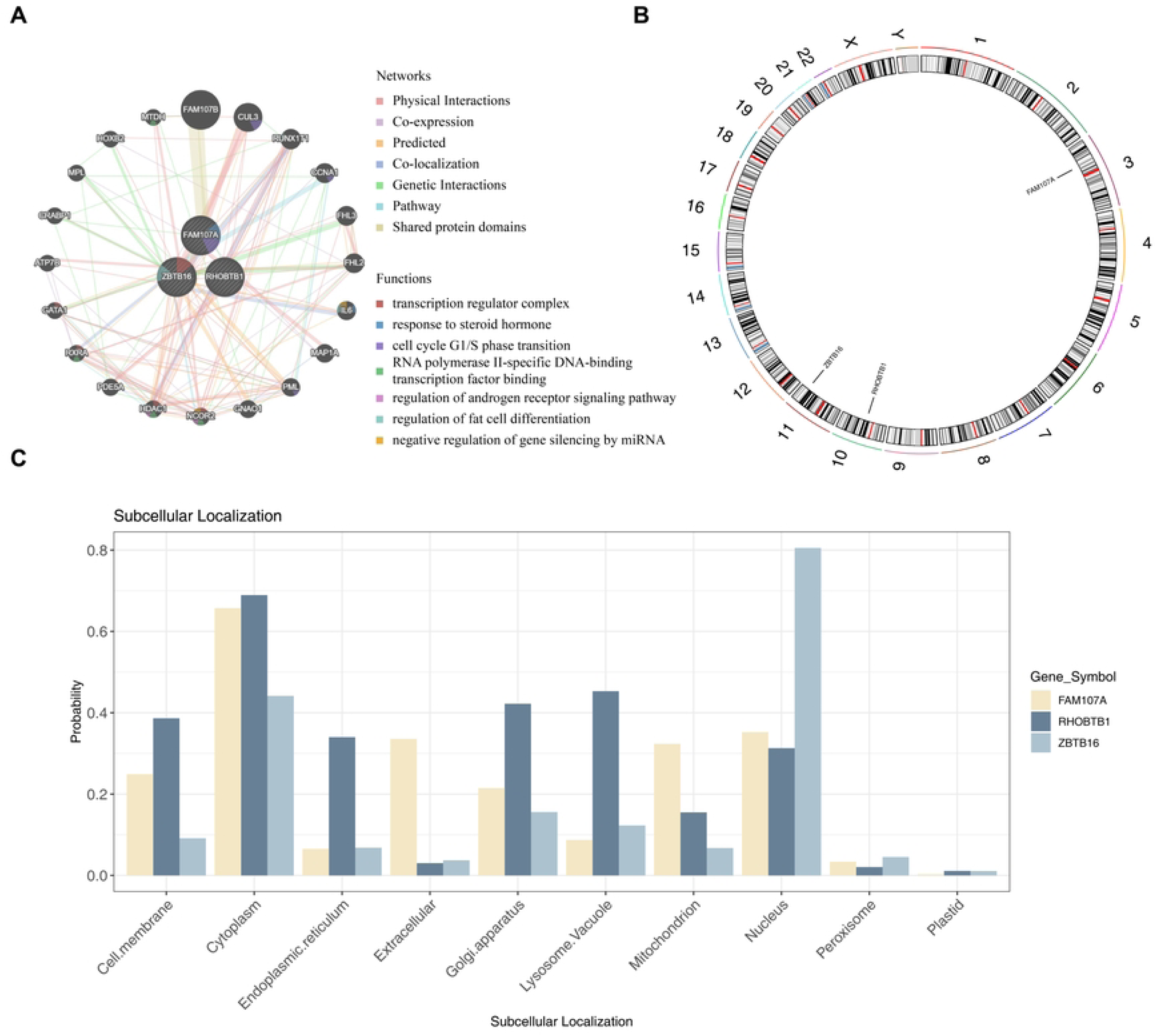
Analysis of genes related to three identified biomarkers. (A) Functional interaction network of the three biomarkers and their predicted functionally-related genes, with color-coded networks and functions including transcription regulator complex and response to steroid hormone. (B) Chromosomal localization map showing the positions of *FAM107A*, *RHOBTB1*, and *ZBTB16*. (C) Subcellular localization probability bar plot for *FAM107A*, *RHOBTB1*, and *ZBTB16*.

### 3.5 Correlation analysis of pathway enrichment and biomarkers

GSEA revealed that *FAM107A* was enriched in 26 KEGG pathways. It showed significant enrichment in pathways such as lysosome, cell adhesion molecules, oxidative phosphorylation, spliceosome, and viral myocarditis (*P* < 0.05, |NES| > 1, FDR< 0.25) (**Fig 5A, S6 Table)**. *RHOBTB1* was found to be enriched in 22 KEGG pathways, with remarkable enrichment particularly in lysosome, and oxidative phosphorylation (FDR< 0.25, |NES| > 1, *P* < 0.05) **(S7 Table, Fig 5B**). *ZBTB16* demonstrated significant enrichment in a total of 15 pathways, including spliceosome, cell adhesion molecules, lysosome, asthma, and allograft rejection (*P* < 0.05, |NES| > 1, FDR< 0.25) **(S8 Table, Fig 5C**). Importantly, among the top 5 pathways, all 3 biomarkers were commonly enriched in the lysosome pathway. These discoveries offered essential understandings regarding the molecular mechanisms that underlie the development of OA.

**Fig 5.**
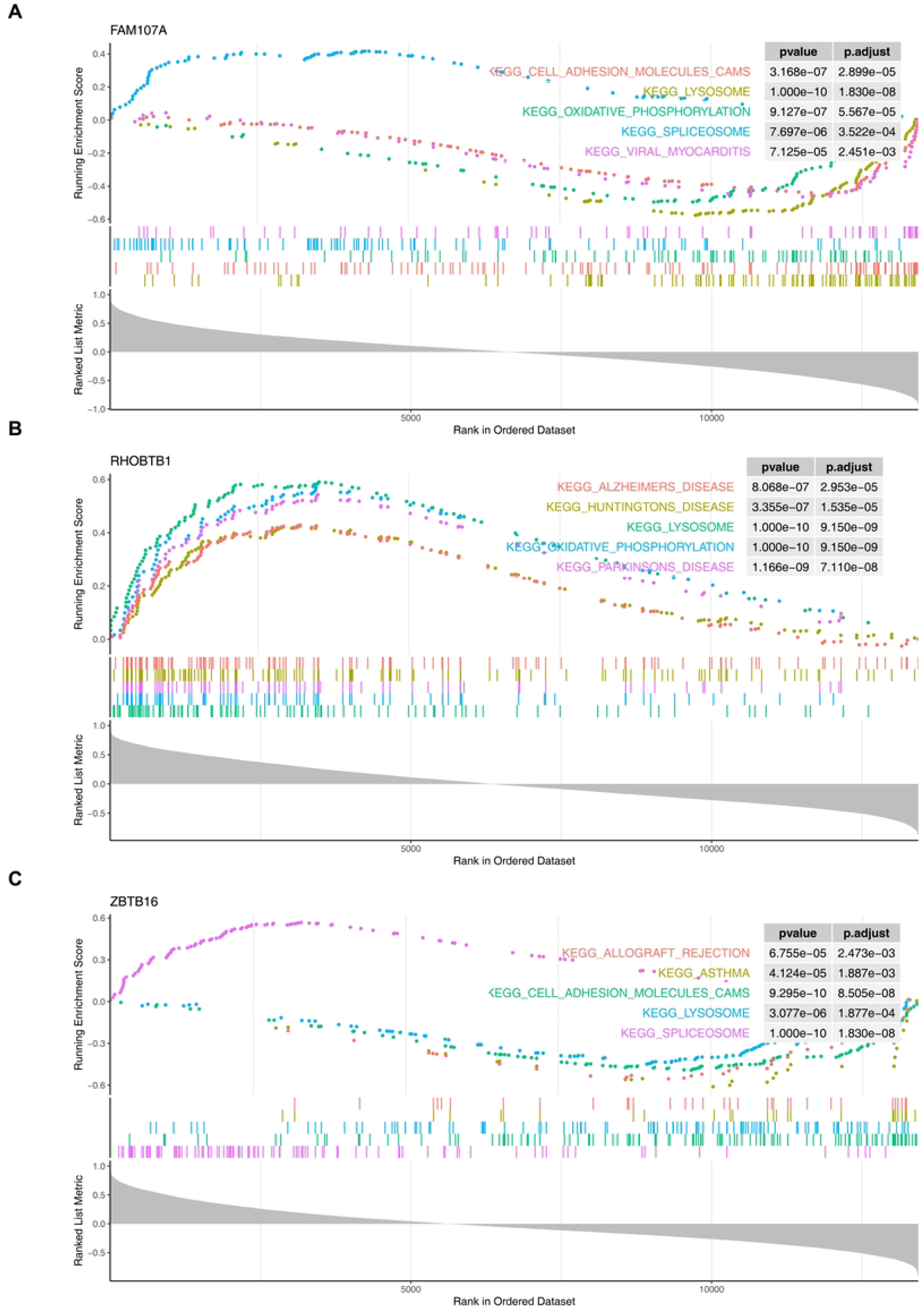
Gene set enrichment analysis (GSEA). (A) GSEA of *FAM107A*. (B) GSEA of *RHOBTB1*. (C) GSEA of *ZBTB16*.

### 3.6 Immune cell profiling and correlation analysis in OA using the CIBERSORT algorithm

The CIBERSORT algorithm was applied to conduct an analysis of the immune cell characteristics within the context of OA. This analysis aimed to explore immune regulation, as well as the correlation between biomarkers and immune cells. A stacked plot depicted the relative abundances and proportional distributions of 22 immune cells in the OA and control individuals of the training set, with varying degrees of infiltration for each cell type (**Fig 6A**). By comparing the differences in immune cells between the OA and control individuals, 6 immune cell types, namely CD8 T cells, activated mast cells, follicular helper T cells, resting mast cells, macrophages M0, and resting memory CD4 T cells, exhibited remarkable differences (*P* < 0.05). Specifically, the proportions of CD8 T cells, activated mast cells, and follicular helper T cells decreased in the OA group, while those of macrophages M0 and resting mast cells increased (**Fig 6B**). Regarding the correlations among these immune cells **(S9 Table, Fig 6C**), activated mast cells and follicular helper T cells demonstrated the most intense positive correlation (r=0.69, *P* < 0.001), and activated mast cells and resting mast cells showed the most pronounced negative correlation (r=-0.86, *P* < 0.001). Further investigation was conducted into the correlations between these immune cells and the 3 biomarkers. The results indicated that CD8 T cells had the greatest positive correlation with *FAM107A* (r=0.72, *P* < 0.001), and activated mast cells had the highest degree of negative correlation with *RHOBTB1* (r=-0.74, *P* < 0.001) **(S10 Table, Fig 6D**).

**Fig 6.**
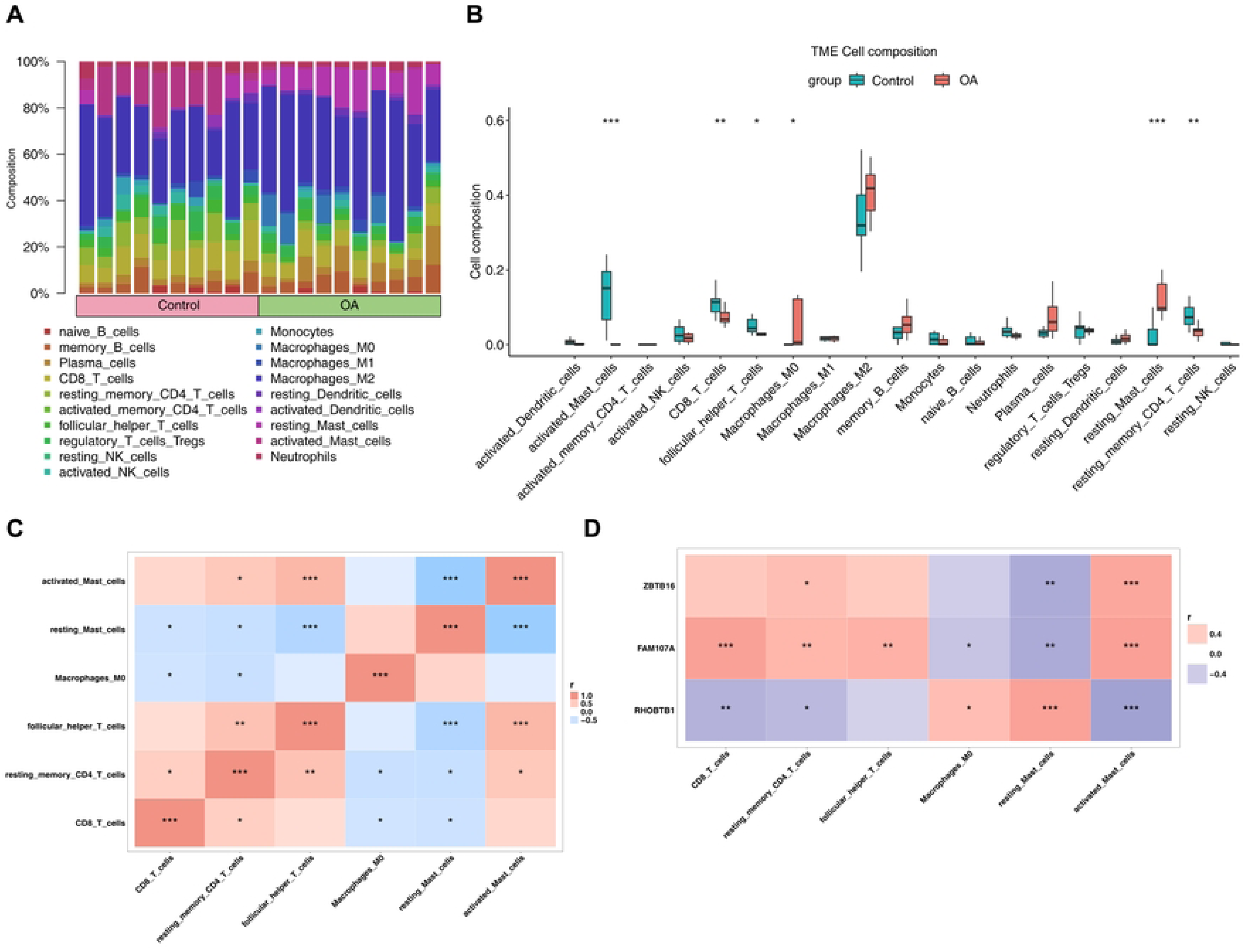
Immune cell profiling and correlation analysis in OA using the CIBERSORT algorithm. (A) Stacked plot illustrating the relative abundances and proportional distributions of 22 immune cell types in OA and control individuals from the training set, showing varying infiltration degrees of each cell type. (B) Box plots comparing the differences in immune cell compositions between OA and control groups, highlighting 6 significantly different cell types with P < 0.05. (C) Correlation heatmap of the significantly different immune cells, depicting their pairwise correlation coefficients and statistical significances. (D) Correlation heatmap between the 3 biomarkers (*FAM107A*, *RHOBTB1*, *ZBTB16*) and key immune cells

### 3.7 Construction of miRNA-mRNA and TF-mRNA regulatory networks

A sum of 76 miRNAs associated with the 3 biomarkers were predicted. Specifically, 27 miRNAs were related to *FAM107A*, 48 to *RHOBTB1*, and 1 to *ZBTB16*. Based on these miRNAs and the 3 biomarkers, a miRNA-mRNA regulatory network was constructed. This network consisted of the 3 biomarkers, 67 miRNAs, and 76 miRNA-mRNA interaction pairs **(S11 Table, Fig 7A**). In the transcription factor-mRNA (TF-mRNA) network, *FAM107A* was regulated by 75 TFs, *RHOBTB1* by 72 TFs, and *ZBTB16* by 442 TFs. By screening the top 30 TFs for 3 biomarkers according to the score, a TF-mRNA regulatory network was established, which included the 3 biomarkers, 21 TFs, and 30 TF-mRNA interaction pairs **(S12 Table, Fig 7B**).

**Fig 7.**
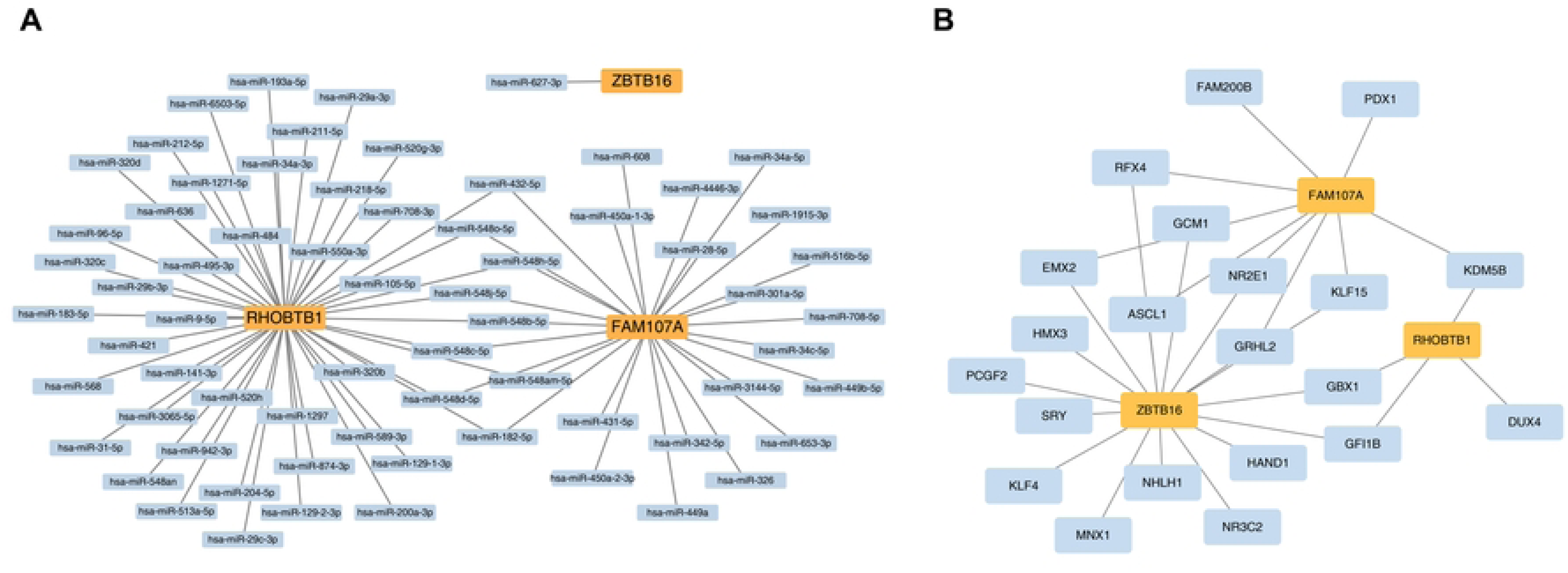
Construction of networks. (A) miRNA-mRNA regulatory network. (B) TF-mRNA regulatory network.

### 3.8 The biomarkers reveal robust binding affinity towards key small molecule compounds intended for targeting

Based on the CTD database, a sum of 100 small molecule compounds targeting *FAM107A* and 112 targeting *RHOBTB1* were predicted. For both *FAM107A* and *RHOBTB1*, bisphenol A, valproic acid, and 4-(5-benzo(1,3)dioxol-5 yl-4-pyridin-2-yl-1H-imidazol-2-yl) benzamide were ranked among top3 in terms of interaction count **(S13-14 Table, respectively)**. Additionally, the 168 small molecule compounds targeting *ZBTB16* were predicted, with benzo(a)pyrene, valproic acid, and tetrachlorodibenzodioxin ranking in the top3 **(S15 Table)**. A small molecule compound-gene interaction network (**Fig 8A**) was constructed, which comprised 3 biomarkers, 40 small molecule compounds, and 60 interaction pairs. The construction was carried out using the top 20 potential small molecule compounds ranked by interaction count for these three biomarkers. Additionally, the binding of the top3 small molecule compounds ranked by interaction count for *FAM107A*, *RHOBTB1*, and *ZBTB16*, respectively, was evaluated (**Table 1-3**, **Fig 8B-J**). The findings indicated that the strongest binding of *FAM107A* and *RHOBTB1* was with 4-(5-benzo (1,3) dioxol-5-yl-4-pyridin-2-yl-1H-imidazol-2-yl) benzamide, with binding energies of -5.99 kcal/mol and -7.58 kcal/mol, respectively. The strongest binding of *ZBTB16* was with benzo(a)pyrene and tetrachlorodibenzodioxin, both with a binding energy of -6.02 kcal/mol. These findings further validated the small molecule compounds’ prediction results.

**Fig 8.**
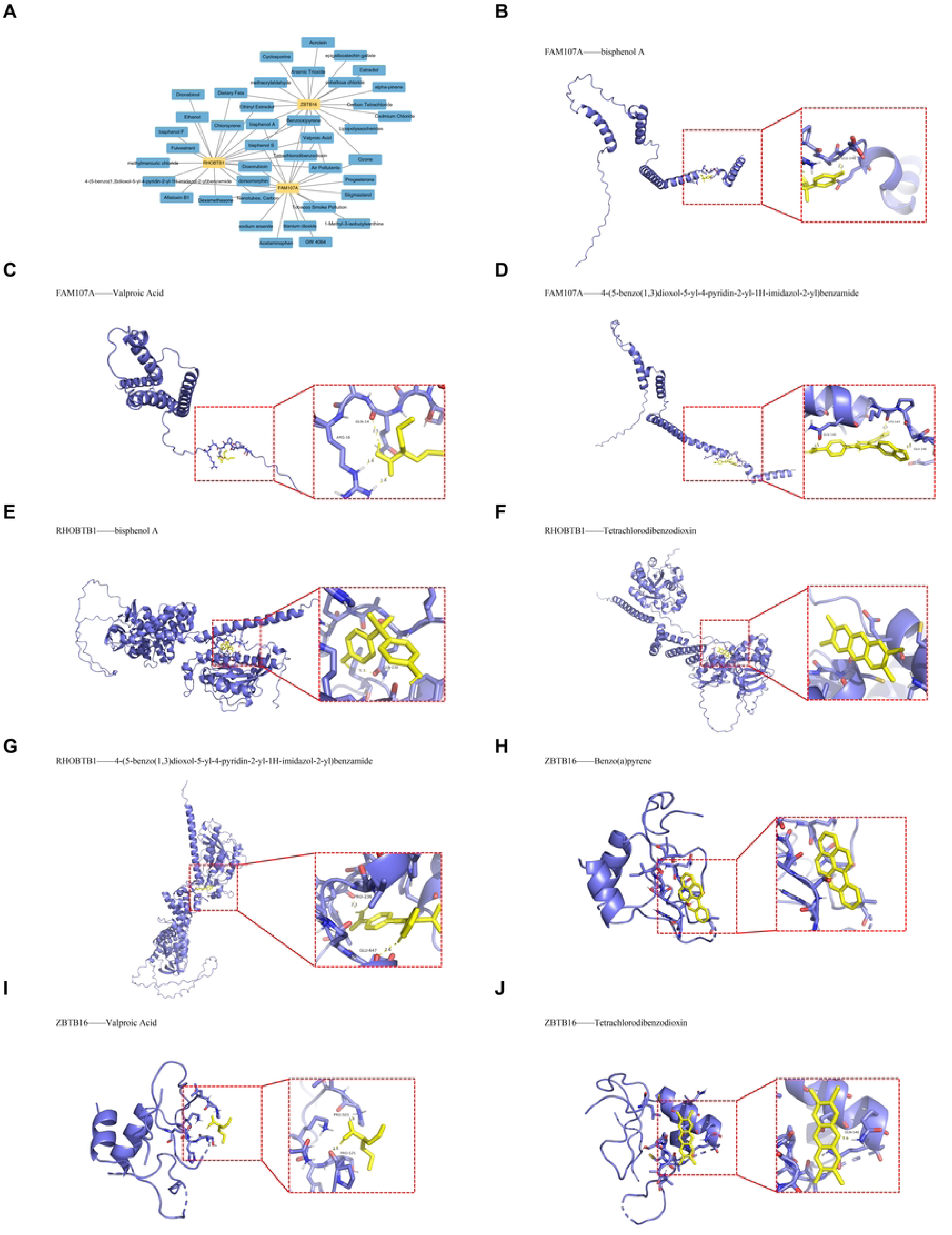
Small molecule compound-gene interaction network and molecular docking analysis. (A) Small molecule compound-gene interaction network consisting of 3 biomarkers (*FAM107A*, *RHOBTB1*, *ZBTB16*), 40 small molecule compounds, and 60 interaction pairs, constructed using the top 20 potential small molecule compounds ranked by interaction count for each biomarker. (B) Molecular docking visualization of *FAM107A* with bisphenol A. (C) Molecular docking visualization of *FAM107A* with valproic acid. (D) Molecular docking visualization of *FAM107A* with 4-(5-benzo(1,3)dioxol-5-yl-4-pyridin-2-yl-1H-imidazol-2-yl)benzamide. (E) Molecular docking visualization of *RHOBTB1* with bisphenol A. (F) Molecular docking visualization of *RHOBTB1* with tetrachlorodibenzodioxin. (G) Molecular docking visualization of *RHOBTB1*with 4-(5-benzo(1,3)dioxol-5-yl-4-pyridin-2-yl-1H-imidazol-2-yl)benzamide. (H) Molecular docking visualization of *ZBTB16* with benzo(a)pyrene. (I) Molecular docking visualization of *ZBTB16* with valproic acid. (J) Molecular docking visualization of *ZBTB16* with tetrachlorodibenzodioxin.

**Table 1.** Binding energy of FAM107A with small molecule compounds.

| gene_name | drug_name | cid | Best_Conformation | Affinity (kcal/mol) |
| --- | --- | --- | --- | --- |
| FAM107A | bisphenol A | 6623 | 29/50 | -4.31 |
| FAM107A | Valproic Acid | 3121 | 33/50 | -2.99 |
| FAM107A | 4-(5-benzo(1,3)dioxol-5-yl-4-pyridin-2-yl-1H-imidazol-2-yl)benzamide | 4521392 | 24/50 | -5.99 |

**Table 2.** Binding energy of RHOBTB1 with small molecule compounds.

| gene_name | drug_name | cid | Best_Conformation | Affinity (kcal/mol) |
| --- | --- | --- | --- | --- |
| RHOBTB1 | bisphenol A | 6623 | 24/50 | -5.39 |
| RHOBTB1 | Tetrachlorodibenzodioxin | 3121 | 40/50 | -6.70 |
| RHOBTB1 | 4-(5-benzo(1,3)dioxol-5-yl-4-pyridin-2-yl-1H-imidazol-2-yl)benzamide | 4521392 | 50/50 | -7.58 |

**Table 3.**
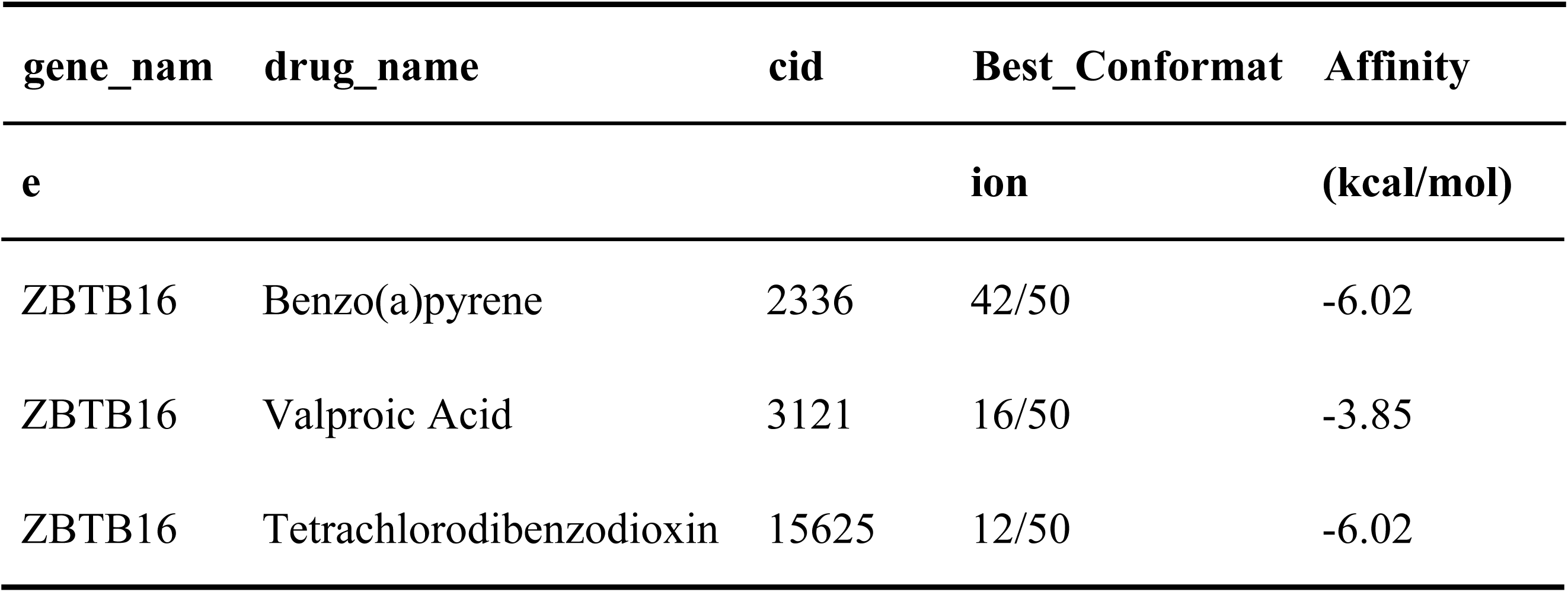
Binding energy of ZBTB16 with small molecule compounds. gene_nam drug_name cid Best_Conformat Affinity.

### 3.9 Validation of biomarker expression in clinical samples

To further confirm the biomarker expression within clinical samples, RT-qPCR was performed on samples from the OA and control individuals. The results demonstrated that, in contrast to the control individuals, the expression levels of *RHOBTB1* (*P* = 0.0023) and *ZBTB16* (*P* = 0.0448) in OA individuals were markedly upregulated (**Fig 9A-B**), while the expression level of *FAM107A* (*P* = 0.0051) was significantly downregulated (**Fig 9C**). These findings support the potential of *RHOBTB1*, *FAM107A*, and *ZBTB16* as biomarkers for clinical assessment of OA.

**Fig 9.**
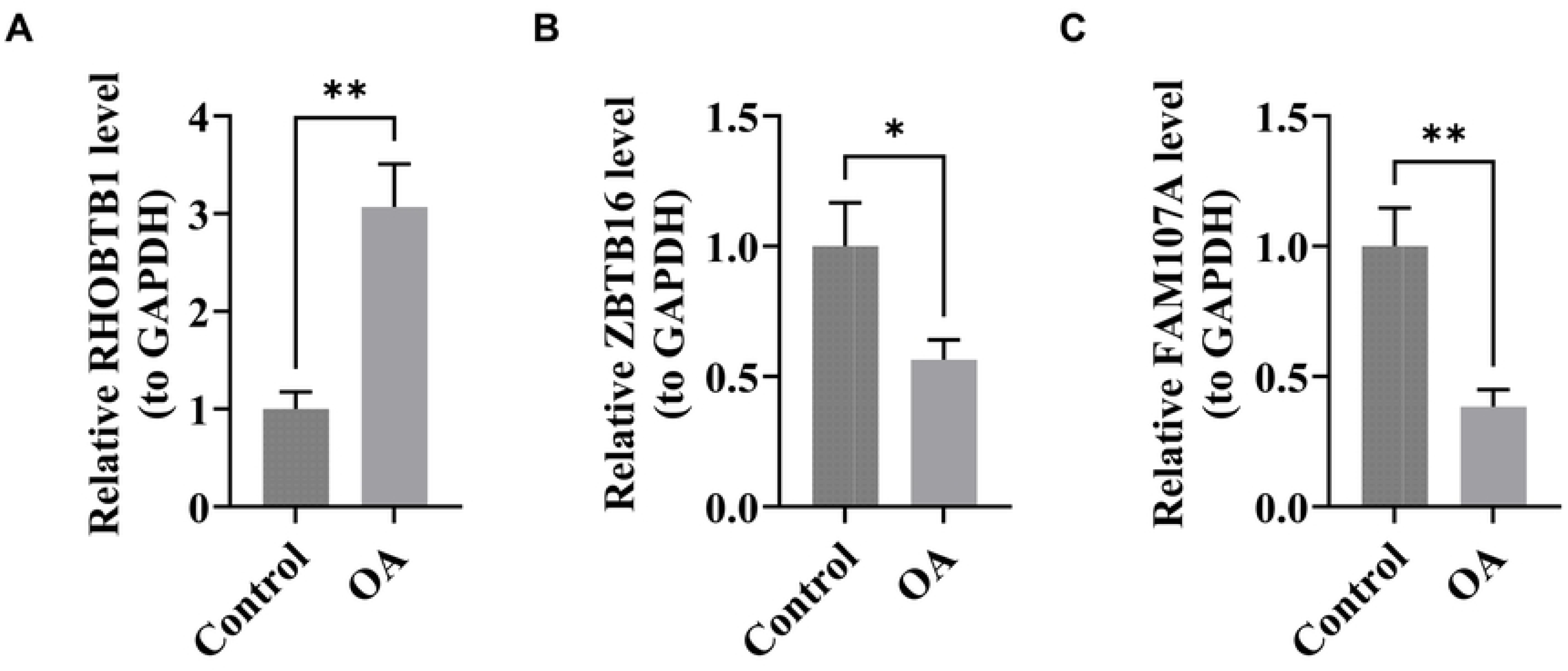
Validation of biomarker expression in clinical samples via RT-qPCR. (A) Bar plot showing the relative expression level of *RHOBTB1* in OA and control clinical samples. (B) Bar plot illustrating the relative expression level of *ZBTB16* in OA and control clinical samples. (C) Bar plot depicting the relative expression level of *FAM107A* in OA and control clinical samples.

## 4 Discussion

Previous studies have emphasized that OA is not merely a “wear and tear” disease but also involves immune dysregulation and metabolic imbalance[3, 5]. DR, as a well-recognized intervention to delay aging and chronic diseases, modulates pathways such as mTOR, AMPK, and sirtuins, thereby reducing oxidative stress and inflammation[10]. In this study, we identified three dietary restriction (DR)-related biomarkers, *FAM107A*, *RHOBTB1*, and *ZBTB16*, in OA through transcriptomic and bioinformatics analyses. These biomarkers were closely associated with immune infiltration and key metabolic pathways, suggesting their potential involvement in OA pathogenesis. Our findings indicate that DR-related genes may link metabolic regulation with immune responses in OA, providing new insights into early diagnosis and therapeutic strategies.

*FAM107A*, located at chromosome 3p21.1, is widely recognized as a tumor suppressor gene and is generally downregulated in various malignancies, often due to promoter hypermethylation. Functionally, *FAM107A* is an actin-associated, stress-inducible protein that modulates F-actin dynamics and thereby participates in cytoskeletal organization and phenotype regulation of cells[28]. Mechanistic studies have revealed that *FAM107A* promotes actin polymerization and regulates key oncogenic processes such as proliferation and migration through the FAK/PI3K/AKT signaling pathway. Importantly, cytoskeletal remodeling, particularly reorganization of filamentous actin, is a hallmark of chondrocyte dedifferentiation and OA progression, linking *FAM107A* dysregulation with OA pathophysiology[29]. These findings highlight that while *FAM107A* has been studied primarily in cancer, its cytoskeletal regulatory functions may provide new insight into mechanisms underlying OA development and progression. In OA, dysregulated immune responses and metabolic reprogramming are also central to disease pathogenesis. Recent transcriptomic studies have revealed that synovial mast cells release histamine, proteases, and inflammatory mediators that exacerbate joint inflammation and cartilage damage[30]. Integrating our findings with this evidence, *FAM107A* may not only be involved in metabolic and stress regulation but also influence immune cell infiltration and inflammatory signaling in OA. Thus, *FAM107A* represents a promising DR-related molecule in OA, with its potential mechanisms likely involving the crosstalk between metabolic control and immune modulation.

*RHOBTB1* is an atypical member of the Rho GTPase family, characterized by a distinct Rho domain and two BTB domains. Unlike classical Rho GTPases, *RHOBTB1* regulates cytoskeletal dynamics through multiple mechanisms, including interaction with ROCK1 to suppress cell invasion and functioning as a substrate adaptor for the Cullin-3 ubiquitin ligase complex to modulate protein turnover. Importantly, *RHOBTB1* directly influences actin polymerization, thereby playing a crucial role in maintaining cellular structural integrity. Recent machine learning analyses of OA synovial tissues have identified *RHOBTB1* as a central regulatory gene, with expression negatively correlated with disease progression and strongly associated with immune cell infiltration, suggesting a protective role[31]. In contrast, our PCR validation in OA models revealed paradoxical upregulation of *RHOBTB1* expression, which may reflect a compensatory regulatory mechanism. Given that cytoskeletal reorganization and protein homeostasis imbalance drive OA pathogenesis through abnormal actin dynamics and chondrocyte dedifferentiation[32], the observed *RHOBTB1* upregulation may represent an adaptive response to reinforce cytoskeletal stability and protein quality control. This suggests *RHOBTB1* plays a dual role in OA: contributing to pathological processes while simultaneously functioning as an intrinsic protective mechanism.

*ZBTB16*, also known as promyelocytic leukemia zinc finger protein, is a multifunctional transcription factor involved in diverse biological processes including stem cell maintenance, immune regulation, and metabolic control[33]. It has been implicated in adipogenesis, glucose and lipid metabolism, cardiac hypertrophy, and fibrosis, thereby serving as a pleiotropic regulator of metabolic syndrome–related traits[33]. In the cardiovascular system, studies have shown that *ZBTB16* upregulation in atrial tissues of type 2 diabetic mice increases atrial fibrillation susceptibility by activating the Txnip-Trx2 axis, leading to mitochondrial oxidative stress and calcium homeostasis imbalance[34].

In the skeletal system, *ZBTB16* plays a crucial role in maintaining tissue homeostasis. Super-enhancer–driven *ZBTB16* expression has been reported to promote osteogenic differentiation of mesenchymal stem cells and protect against osteoporosis, underscoring its importance in skeletal homeostasis[35]. This function is highly relevant to understanding its role in OA, a disease characterized not only by cartilage degradation but also by subchondral bone remodeling and skeletal homeostasis disruption. However, *ZBTB16* expression and function in OA appear to be complex. Previous studies reported its downregulation in synovial and cartilage tissues of OA patients[36, 37], while functional experiments demonstrated that *ZBTB16* overexpression attenuates lipopolysaccharide-induced chondrocyte apoptosis, inflammatory cytokine release, and extracellular matrix degradation, partly through transcriptional repression of GRK2, a kinase that accelerates cartilage degeneration[36]. In contrast, PCR results from this study showed significantly increased *ZBTB16* expression in OA patients, suggesting potential tissue-specific, stage-dependent, or compensatory regulatory differences during disease progression. Bioinformatics and machine learning analyses further support *ZBTB16* as a central diagnostic biomarker for OA, highlighting its close association with immune cell infiltration, particularly T cells and macrophages that drive synovial inflammation[37].

Taken together, *ZBTB16* exhibits a complex regulatory network in skeletal homeostasis and OA pathogenesis, simultaneously promoting protective osteogenic differentiation and modulating inflammatory cascades. The observed discrepancies in its expression across studies emphasize the importance of clarifying its precise mechanistic roles in different OA tissues and disease stages. Such understanding may provide significant translational value for developing diagnostic strategies and therapeutic interventions targeting bone–cartilage homeostasis.

Our GSEA results show that three core genes are co-enriched in lysosomal pathways, indicating lysosomal dysfunction as a key mechanism in OA pathogenesis. This finding aligns with current OA research frontiers. TFEB dysregulation is central to lysosomal dysfunction. Studies show ox-LDL impairs TFEB activity, inhibits autophagy, and accelerates chondrocyte death and cartilage degradation. Restoring TFEB activity through rapamycin or statins rescues lysosomal function, supporting its role in OA progression[38]. Lysosomal membrane destabilization is another critical mechanism. Hydroxyapatite crystals in OA cartilage induce lysosomal membrane permeabilization, cathepsin B release, NLRP3 inflammasome activation, and pyroptosis[39], consistent with our immune pathway enrichment results. Lysosomes integrate mitochondrial autophagy and ferroptosis networks. When lysosomal function is impaired, mitochondrial quality control fails, leading to oxidative stress, ECM catabolism, and chondrocyte death[40]. RNA networks also connect to lysosomal function. Engineered exosomes delivering miR-140 suppress catabolic proteases and ameliorate OA progression, potentially through regulating lysosomal gene expression[41].

In summary, our GSEA-identified lysosomal pathway integrates multiple aspects of chondrocyte immunometabolic remodeling: TFEB regulation, membrane stability, mitochondrial quality control, and RNA networks. This validates our biomarkers and reveals lysosomes as promising therapeutic targets—including TFEB activators, membrane stabilizers, mitochondrial protectants, and miRNA-based therapies [40–43]. This study compared immune cell differences between OA and control groups through immune infiltration analysis and found significant differences in six types of immune cells: CD8+ T cells, activated mast cells, follicular helper T cells, resting mast cells, M0 macrophages, and resting memory CD4+ T cells. M0 macrophages showed significant differences, which may reflect early changes in the macrophage polarization process. Consistent with recent bioinformatics research findings, this abnormality in M0 macrophages may further lead to significant enrichment of macrophages (particularly the M1 phenotype) in osteoarthritic tissues[42], suggesting a dynamic transition process from resting state to pro-inflammatory polarization. Meanwhile, protective cell subsets such as CD8+ naive T cells and natural killer T (NKT) cells were reduced[42]. This imbalance suggests a shift toward a pro-inflammatory state, driving persistent synovitis and cartilage degradation.

Among adaptive immune cells, CD8⁺ T cells emerged as a key population. Clinical analyses have demonstrated that CD8⁺ T cells in OA joints differentiate into Tc1 (IFN-γ⁺) and Tc17 (IL-17A⁺) subsets, which are markedly elevated in synovial fluid and synovial membrane compared to peripheral blood[43]. These effector subsets are potent drivers of inflammation, matrix catabolism, and pain, indicating their central role in OA progression. Innate immunity also contributes substantially. Mast cells, long overlooked in OA, are increasingly recognized as pathogenic. Transcriptomic and proteomic studies show their accumulation in osteophytes and synovial fluid, where they differentiate under the influence of the OA microenvironment and secrete tryptase, chymase, CPA3, and histamine[30, 44]. These mediators not only degrade extracellular matrix components but also amplify synovitis through activation of fibroblasts, macrophages, and endothelial cells. Moreover, experimental blockade of histamine H1 receptors reduced OA severity in murine models, underscoring the translational relevance of mast cell activity[30]. Recent reviews further emphasize the multifaceted roles of mast cells, which can exert both pro- and anti-inflammatory effects depending on activation pathways. While IgE/FcεRI-driven activation promotes cartilage destruction, alternative phenotypes may modulate inflammation under certain contexts[45]. This duality highlights the need for careful characterization of mast cell subsets in OA.

Integrating our results with these findings, the observed immune infiltration profile—dominated by M1 macrophages, mast cells, and pro-inflammatory CD8⁺ T cell subsets—supports the concept of OA as an immuno-inflammatory disease rather than a purely degenerative disorder. Our data suggest that therapeutic strategies targeting immune pathways, such as modulation of CD8⁺ T cell subsets, rebalancing macrophage polarization, or inhibiting mast cell mediators, could provide novel avenues for disease modification.

Our drug-gene interaction analysis identified candidate small-molecule compounds that may modulate OA progression. Valproic acid (VPA) emerged as a promising therapeutic candidate, targeting hub genes *FAM107A*, *RHOBTB1*, and *ZBTB16*. As an HDAC inhibitor, VPA suppresses matrix-degrading enzymes like MMP13, protects chondrocytes from apoptosis, and activates protective PI3K-AKT signaling pathways[46].

Conversely, we identified harmful environmental compounds, including tetrachlorodibenzodioxin (TCDD) and benzo[a]pyrene (B[a]P). TCDD, an aryl hydrocarbon receptor (AhR) ligand, activates NF-κB and ERK pathways, upregulating pro-inflammatory cytokines (IL-1β, IL-6, IL-8) and exacerbating synovial inflammation[47]. Similarly, B[a]P accelerates bone loss and promotes chondrocyte apoptosis through AhR/Cyp1a1 signaling[48]. Our docking results confirmed strong TCDD binding to matrix-degrading genes (*MMP3*, *BMP2*). These compounds exhibit contradictory metabolic effects compared to beneficial dietary restriction. While TCDD causes pathological weight loss (∼30% in 10 days) through acute toxicity rather than healthy caloric restriction[49, 50], VPA shows obesogenic effects (+0.49 kg)[51]. Notably, dietary restriction paradoxically increased B[a]P-DNA adduct formation by 22% and enhanced cytochrome P450 activity, suggesting enhanced carcinogen toxicity under restricted feeding[52]. VPA shows promise as a disease-modifying OA treatment, while TCDD and B[a]P represent environmental threats to joint health, highlighting the dual nature of small molecules in OA pathogenesis and therapy.

In summary, our study comprehensively explored the interplay between dietary restriction–related genes and osteoarthritis through integrated bioinformatics, immune infiltration profiling, GSEA enrichment, and small-molecule compound prediction. We identified three potential biomarkers (*FAM107A*, *RHOBTB1*, and *ZBTB16*) that are functionally linked to metabolic regulation, immune modulation, and skeletal homeostasis. Moreover, our analyses revealed critical pathogenic pathways, including autophagy–lysosome dysfunction, ferroptosis–mitophagy imbalance, and inflammasome activation, and highlighted VPA as a promising candidate for disease modification, while underscoring the detrimental effects of environmental toxins such as TCDD and benzo[a]pyrene. Collectively, these findings provide novel insights into OA pathogenesis and suggest potential targets for early diagnosis and therapeutic intervention.

Nevertheless, several limitations should be acknowledged. First, the transcriptomic data were derived from public databases with limited sample sizes, which may introduce bias and reduce generalizability. Second, our results were mainly based on in silico analyses, and functional experiments—although partially validated—remain insufficient to fully elucidate the molecular mechanisms. Third, while drug–gene docking offered valuable predictions, in vitro and in vivo validation of compound efficacy and safety in OA models is still required. Future studies integrating multi-omics data, larger clinical cohorts, and experimental validation will be necessary to strengthen the translational relevance of our findings.

## 5 Conclusion

In this study, we identified three DR-related biomarkers in OA, namely *FAM107A*, *RHOBTB1*, and *ZBTB16*, by integrating transcriptomic data from public databases with multiple bioinformatics analyses. Functional characterization revealed their involvement in gene–gene interaction networks, chromosomal distribution, subcellular localization, biological pathways, and immune cell infiltration, suggesting potential roles in OA pathogenesis. Furthermore, several candidate small-molecule compounds targeting these biomarkers were predicted and validated through molecular docking, offering novel therapeutic opportunities. The expression of these biomarkers was further confirmed in clinical samples. Collectively, our findings provide a comprehensive insight into the molecular mechanisms of DR-related biomarkers in OA and highlight their potential as targets for precision therapy.

## Data Availability

The datasets analysed in this study are available in the Gene Expression Omnibus (GEO) database (http://www.ncbi.nlm.nih.gov/geo/), including GSE89408 dataset.

## Acknowledgements

We would like to express our sincere gratitude to all the individuals and organizations who supported and assisted us throughout this research. Special thanks go to the contributing authors for their valuable efforts and collaboration. Finally, we extend our heartfelt appreciation to everyone who has accompanied and encouraged us along the way — this work would not have been possible without your support.

## Author contributions

SW: Investigation, Methodology, Software, Project administration, Writing-original draft, Writing-review and editing.

XL: Investigation, Software, Data curation, Visualization, Writing-original draft. ZX: Visualization, Validation, Writing-original draft.

JW: Supervision, Project administration, Resources, Writing-review and editing.

This work currently described has not been published, is not being considered for publication elsewhere, and its publication was approved by all authors.

## Ethics approval and consent to participate

The study was conducted in accordance with the Declaration of Helsinki. The research study has been approved by the Ethics Committee of Nanjing Drum Tower Hospital, Affiliated Hospital of Medical School, Nanjing University. The approval number and date of approval are as follows: [2025-0615-01] and [August 13th, 2025]. Participants in this study were provided with a clear and understandable explanation of the research objectives, procedures, potential risks, and benefits. They were informed that their participation is voluntary and that they have the right to withdraw from the study at any time. Participants were given the opportunity to ask questions and provided written informed consent prior to their involvement in the study.

## Consent for publication

Not applicable.

## Supporting information captions

**S1 Table** A total of 276 dietary restriction-related genes (DRRGs) were obtained in this study.

**S2 Table** qPCR primer sequences.

**S3 Table** The 43 candidate genes identified in this study.

**S4 Table** GO enrichment analysis.

**S5 Table** KEGG enrichment analysis.

**S6 Table** GSEA of *FAM107A*.

**S7 Table** GSEA of *RHOBTB1*.

**S8 Table** GSEA of *ZBTB16*.

**S9 Table** Correlation analysis of differential immune cells.

**S10 Table** Correlation between biomarkers and differentially infiltrating immune cells.

**S11 Table** miRNAs with potential interactions with biomarkers.

**S12 Table** TFs with potential regulatory effects on biomarkers.

**S13 Table** Drugs predicted to target *FAM107A*.

**S14 Table** Drugs predicted to target *RHOBTB1*.

**S15 Table** Drugs predicted to target *ZBTB16*.

